# Physiotherapy service during the COVID-19 pandemic in Nepal: An onsite survey and the lived experience among clinicians

**DOI:** 10.64898/2026.03.19.26348776

**Authors:** Nishchal Ratna Shakya, Samiksha Dahal, Nistha Shrestha, Gillian Webb, Ann-Katrin Stensdotter

**Affiliations:** Department of Physiotherapy, Kathmandu University School of Medical Sciences, Dhulikhel, Kavre, Nepal; Faculty of Medicine and Health Sciences, Department of Neuromedicine and Movement Science, Norwegian University of Science and Technology (NTNU), Trondheim, Norway; Epidemiology and Disease control division, Department of health services, Ministry of Health and Population, Government of Nepal; Faculty of Medicine, Dentistry and Health Sciences, The University of Melbourne, Australia

**Keywords:** COVID-19, Nepal, Physiotherapy, Health service delivery, Rehabilitation workforce, Frontline healthcare providers, Low– and middle-income countries

## Abstract

**Background:** The COVID-19 pandemic significantly disrupted healthcare services globally, particularly in low-resource settings. This study explores the impact of the pandemic on physiotherapy services in Nepal.

**Methods:** A cross-sectional study was conducted. Qualitative data were collected through semi-structured interviews with 12 physiotherapists, while quantitative data were gathered from an onsite survey of 29 health facilities at six different districts of Province III of Nepal. Inductive thematic analysis approach was used to analyze the qualitative data, and descriptive statistics were used for the closed ended questions.

**Results:** The findings were categorized into sub-themes under two major themes: i) Pandemic effect on physiotherapy services and patient care and ii) Adaptation, innovation and collaboration. The study revealed a significant disruption in physiotherapy services with a notable decline in patient flow and service availability. Most patients, especially those with disabilities and post-operative needs, experienced worsening conditions due to limited access to care. There was an increased recognition of the role of physiotherapy in acute respiratory care and post-COVID-19 recovery. Tele-rehabilitation was explored as an alternative care method but faced challenges in implementation. More than half (62.07%) of the centers reported uninterrupted physiotherapy services, whereas almost one third (31.03%) experienced service suspension. Most centers (89.7%) had personal protective equipment available, and majority (86.2%) of the physiotherapists worked in multidisciplinary team: fever clinics, triage, emergency care, respiratory physical therapy, and nursing and administrative support were among the expanded roles. Several centers (37.9%) used virtual care with telephone consultation serving as the primary modality. Virtual service was mostly absent in centers where in-person services persisted.

**Conclusion:** The COVID-19 pandemic significantly impacted physiotherapy services in Nepal, leading to service disruptions and compromised patient care. It highlighted the need to further incorporate physiotherapy into the healthcare system and enhance rehabilitation services to improve continued patient care.

## Introduction

The coronavirus disease 2019 (COVID-19) was first detected in Wuhan, China in December 2019. Following its widespread community transmission, World Health Organization (WHO) declared it a pandemic on 11 March 2020 [1]. World Physiotherapy stated 87% of its member organizations reported service disruptions during this period [2]. Globally, approximately 2.4 billion people are estimated to require rehabilitation services, with more than half of these needs remaining unmet in low– and middle-income countries (LMICs). This situation was further exacerbated by the pandemic [3]. The pandemic placed an immense strain on the already limited rehabilitation capacity posing a threat to many LMICs healthcare models which are fragile and dependent on international funds to provide routine care [4].

In Nepal, a government-imposed lockdown and institutional restrictions for national preparedness activities aimed to decrease community transmission, impacting healthcare services for both COVID-19 and non-COVID-19 patients [5]. Fear of infection caused reluctance in seeking medical care, while closure of outpatient departments (OPDs), staff shortages and delay in treatment further hindered access to essential health services [6], including physiotherapy.

Physiotherapy, an established profession throughout the world, plays a vital role in developing, maintaining and improving physical function and movement, thereby improving quality of life [7]. In Nepal, physiotherapists who work in primary healthcare facilities are involved in the management of patients admitted to hospital. This includes contagious infections such as COVID-19. Physiotherapy plays a key role in physical rehabilitation including respiratory management, therefore it was central in the care of patients with COVID-19 [8]. A number of health facilities in Nepal launched post COVID rehabilitation center [9,10] and employed dedicated staff. World Physiotherapy noted that individuals with long-term conditions were particularly affected by the disruption in physiotherapy services, as they were more susceptible to complications related to their pre-existing conditions [2]. People with existing disabilities (PWDs) were significantly affected by the COVID-19 pandemic, facing challenges in access to healthcare services [11]. A study conducted in 35 European countries revealed that nearly 2.2 million people affected by disability were severely impacted by reduced or no access to rehabilitation services due to the pandemic [12]. Considering the availability, access and coverage of physiotherapy in Europe compared to LMIC countries in general, we considered it important to examine the situation in Nepal during the COVID pandemic. Hence this study was aimed to explore the impact of COVID-19 on physiotherapy services in Nepal.

## Methods

### Study design

This cross-sectional study, based on interviews and surveys, was part of a larger study about physiotherapy service in Nepal [13], undertaken in the province III during the ongoing pandemic in 2020–2022. Province III is the second most populated area including the capital Kathmandu. Narrative data was collected via online interviews with physiotherapists about their lived experiences from working during the pandemic. Consolidated criteria for reporting qualitative research (COREQ) was followed [14] (Detail in S1 File). An onsite health facility survey was conducted on the status of service provided by physiotherapists during pandemic.

### Recruitment and selection

Interviews with physiotherapists were conducted between June 2020 and May 2021 during the lockdown. Inclusion criteria was a minimum of one year of working experience and having knowledge about and experience with physiotherapy services in Nepal. Participants were recruited for interviews by a purposive sampling method [15] to ensure variability related to workplace setting, with the aim of increasing the depth of understanding and retrieving useful information [16]. It was proceeded until saturation of data when no new and significant information was received [17]. The health facility survey was conducted between September 2021 and February 2022. Only healthcare service centers providing physiotherapy services were selected. A stratified purposive sampling method was used for health facilities selection, with the aim of including all levels of the health care system.

### Ethical considerations and data security

Approvals were granted by the Nepal Health Research Council (NHRC-ERB Protocol No. 455/19), the Institutional Review Committee of Kathmandu University School of Medical Sciences / Dhulikhel Hospital (IRC-KUSMS 281/19, 213/24), and the Norwegian Centre for Research Data (NSD 383963). Participants received verbal and written information. Written informed consent was obtained from the participants of interview and health facility survey. Participation was voluntary and participants were free to withdraw from the study at any time. Survey responses were anonymous, and without person identifiable data. Handling of personal data followed the EU General Data Protection Regulation (GDPR) and Norwegian center for research data (NSD) regulations on privacy policy. The study was performed in accordance with the Helsinki Declaration [18].

### Data acquisition

Due to pandemic restrictions, all interviews were performed online in Nepali language using the university licensed Zoom compliant with GDPR (https://explore.zoom.us/en/gdpr/). The questionnaire was constructed by adapting questions to the Nepali contexts from “COVID-19 Quick Canadian Physiotherapist Survey Questionnaire” [19,20] and an online questionnaire provided by the World Physiotherapy [2]. Three experts in physiotherapy with academic, clinical and public health background, discussed and modified the interview and health facility survey questions. The interview guide was piloted with three physiotherapists from a University hospital, rehabilitation and clinical settings. The final interview guide included questions about; how physiotherapy services were managed during COVID; how existing health conditions were affected due to the pandemic in patients requiring physiotherapy or rehabilitation; what alternative strategies were used in treatments and how a change in perception about physiotherapy was understood by other health professionals (S2 File). The facility survey included questions on the general status of physiotherapy services during COVID; use or availability of personal protective equipment (PPE); the role of physiotherapy and questions on virtual care practices (S3 File). The survey questions were selected based on contextual relevance in Nepal. It was intended to capture baseline service disruptions, capacity and adaptability of care delivery. The aim was to focus on profession-specific details about PPE availability, rather than broader hospital PPE shortages, and to investigate how well (or poorly) facilities protected staff providing hands-on care. The survey included items designed to identify how physiotherapy services were adapted, such as role expansion, scope modification or redeployment during waves of the pandemic.

### Data analysis

All recorded interviews were transcribed verbatim in Nepali and translated to English. Accuracy of transcriptions, translations and sufficient de-identification of content was secured. We adopted the six steps of inductive thematic analysis that included familiarization with the data, generating initial codes, searching for themes, reviewing themes, defining and naming the themes. We used a 15-point checklist of criteria to generate initial codes that clustered relevant texts into potential categories, sub-themes and themes [21]. The preliminary codes were identified from the initial interviews and a set of selected priori codes were added to develop a codebook [22] which was reviewed among investigators for consensus. The data was organized manually by color coding the interview transcripts that helped in identifying, sorting and grouping related text excerpts into sub-themes and themes.

The health facility survey data were analyzed descriptively using SPSS Version 27.

## RESULTS

### Participants

Table 1 summarizes the demographic and professional characteristics of the interviewed participants, including age, sex distribution, academic qualifications, years of experience and workplace settings. Saturation of data was reached after 12 interviews. Sub-themes and themes identified through the thematic analysis are outlined in Table 2.

**Table 1:** Demographic characteristics of interview participants (N=12)

| Characteristics | N (%) |
| --- | --- |
| Age (Mean/ SD) | 36.08 (9.87) |
| Gender |  |
| Male | 5 (41.67%) |
| Female | 7 (58.33%) |
| Work experience (Mean/ SD) | 8.33 (5.4) |
| Education level |  |
| Bachelors | 5 (41.67%) |
| Masters | 6 (50%) |
| PhD | 1 (8.33%) |
| Setting |  |
| Rehabilitation centers | 4 (33.33%) |
| Corporate hospital | 1 (8.33%) |
| Private secondary hospital | 1 (8.33%) |
| Special school * | 1 (8.33%) |
| Public hospitals | 4 (33.33%) |
| Community hospital | 1 (8.33%) |

\* A dedicated institution offering therapeutic care alongside education to maximize the physical potential of children with disability

**Table 2:** Example quotes and theme generation.

| Example quotes | Categories | Sub-themes | Themes |
| --- | --- | --- | --- |
| “We have been providing services equally in both OPD and IPD.” (P9, Public hospital) | Physiotherapy service availability and utilization | Impact on physiotherapy services and patient care | Pandemic effect on physiotherapy services and patient care |
| “We were assessing each patient individually and scheduled them for follow up sessions if only they needed. Otherwise, we provided them with mainly home exercises. But now, we advise starting with home stretching, and we ask them to visit us only if there's no improvement. Comparatively, patients now receive less direct care and have to adjust.” (P3, Corporate hospital) | Service standards and delivery management |  |  |
| “..... pandemic has definitely affected the patient's psychology. First, they are already bed-bound due to the disability and additionally, the services were abruptly stopped.....They are very prone to develop secondary complications....” (P2, Rehabilitation center) | Patient flow and impact on patients' health |  |  |
| “Number of patients decreased at first. Patients were afraid of hospitals and rehabilitation centers; they think that we could be the source of infection.” (P3, Corporate hospital) | Challenges in providing patient care | Pandemics add up challenges to providing physiotherapy services |  |
| “Due to lack of proper follow up, there were many patients whose knee function deteriorated. .... There were movement restrictions and reductions in functional abilities.” (P7, Government hospital) | Added challenges: Lesser follow up, unawareness, uncooperative medical professionals, stress, fear of infections and physical exertion |  |  |
| “Senior doctors did not refer the patients at all, and told the patients to go home after plaster cast removal, teaching them just two-four exercises. They knew that the patients would get better if they refer to physiotherapists but they didn't do that during COVID time.” (P3, Rehabilitation center) |  |  |  |
| “We have started collecting patients' contact information forming the team and moving towards digital rehabilitation. We are providing exercises remotely. If this continues, we plan to fully implement tele-rehabilitation (P12, Academician) | Tele-rehabilitation | The change in physiotherapy practice and changing perceptions | Adaptation, innovation and collaboration should be promoted for strategic intervention and awareness of physiotherapy services during pandemics |
| “A nurse whose father was infected with COVID-19 asked me for his exercise recommendations.... People have begun to understand that physiotherapy can really help with recovery” (P3, Rehabilitation center) | Change of perception |  |  |
| “..... we as physiotherapists are normally not used to night duty..... Being away from family for weeks and months were our challenges. To ease the situation, we must have good management for the staff like food, logistics arrangement, proper communication, transportation facilities and information sharing. All these things had helped me as a team leader to convince my staff and motivate them.” (P2, Rehabilitation center) | Strategic physiotherapist or team resolution | Strategies for improving and strengthening physiotherapy services |  |
| “While entering our department, we didn't let the patients enter directly. Even for counselling and communication, we |  |  |  |
| *maintained proper distance. We didn't allow any visitors inside without proper sanitization. Washing their hands and wearing masks were compulsorily." (P5, Government disability sector)* | *Patient management strategies* | *during pandemics* |  |
| *"And this pandemic is also an opportunity to raise awareness regarding physiotherapy. Many people, including doctors, nurses and other health workers, were not fully aware of the importance of physiotherapy. .... Even WHO (World Health Organization) had confirmed that physiotherapy helps patients recover and reduce mortality." (P3, Rehabilitation center)* | *Informing the scope and recognition* |  |  |

### Pandemic effect on physiotherapy services and patient care

#### Impact on physiotherapy services and patient care

##### Effect on physiotherapy service availability and utilization

Responses from physiotherapists regarding the status of services during pandemic showed variability. Only a few physiotherapists reported that their institutions were able to provide uninterrupted physiotherapy services and maintain daily services. Some reported complete unavailability of physiotherapy treatment and halted services. It was reported that OPD services were often more impacted than inpatient department (IPD) services. Some reported that physiotherapists provided exercise interventions to COVID patients at the critical care unit.

> *“In our organization, we attended IPD patients daily during the lockdown period. There were seven of us and each of us was assigned a day in a week. The OPD was completely closed for the first two months of lockdown.” (P3, Corporate hospital)*

##### Effect on service standards and delivery management

Physiotherapists reported that the quality and quantity of treatment were hampered due to limitations on in-person visits and resources. While treatments were still provided according to the specific needs of each patient, they were often given on a priority basis with a focus on essential care. Exercise programs, aimed primarily at maintaining patient function, were emphasized, however respondents stated that the overall quality of service was affected. Most of hands-on techniques, such as soft tissue and joint mobilization, were also hindered.

> *“We were assessing each patient individually and scheduled them for follow up sessions if only they needed. Otherwise, we provided them with mainly home exercises.” (P3, Corporate hospital)*

##### Change in patient flow and impact on patients’ health

The majority of physiotherapists reported a noticeable decrease in patient flow with a reduction in both patient visits and admissions. Many physiotherapists indicated that patients experienced considerable psychological distress, especially those with pre-existing disabilities. The impact was reported to be more severe for individuals with pre-existing conditions as they struggled with both the direct and indirect consequences of their health issues. The physiotherapists also reported that pediatrics and neurological patients experienced the greatest challenges with their specialized treatment needs often being unmet due to disruptions in services. Development of secondary symptoms was commonly reported with many patients experiencing a decline in physical function. The physiotherapists observed that improvements were not always as good as expected.

> *“Compared with the number of patients before, it is very low. It is a bit difficult to say whether the patients don’t want to come because of their pain or due to the fear of COVID-19.” (P8, Private secondary hospital)*

> *“Because of COVID-19, many patients were discharged early, and lack of follow-ups affected outcomes significantly. In musculoskeletal cases, especially post-operative conditions during pandemic, we’ve seen patients returning with restricted mobility in follow ups due to lack of physiotherapy.” (P12, Academician)*

#### Pandemics add up challenges to providing physiotherapy services

##### Challenges in providing patient care during pandemic

Physiotherapists pointed out that the lack of transportation facilities hindered the ability of patients to visit physiotherapy centers. They also reported reduced visits due to the patients’ hesitation to seek physiotherapy treatment as they were concerned about contracting illness.

> *“Many children who were discharged from hospital before COVID-19 couldn’t come for follow up. So, the parents contacted us regarding the problems their children were facing and how they could manage by themselves. They stated of having their own obligations and facing issues with transportation.” (P1, Rehabilitation center)*

##### Other challenges: Lesser follow up, unawareness, uncooperative medical professionals, stress, fear of infections and physical exertion

Another major challenge reported by physiotherapists was the lack of follow-ups, which hindered the ability to track patient progress and adjust treatment plans accordingly. Physiotherapists also reported being unaware of government-level initiatives that created uncertainty around proper protocols and support. Some participants also experienced issues with uncooperative medical professionals as they hesitated in referring patients to physiotherapy. The physical and mental stress, experienced by both patients and physiotherapists, was reported by most of the physiotherapists, exacerbated by the need to work in PPE and the difficulty in administering interventions.

> *“I have no idea or discovered anything until now that Nepal government has done in disability sector in coordination with physiotherapists. But I’m sure that the physiotherapists employed in Nepal government hospitals are working in their respective areas with utmost caution and cautiously providing the continuous care.” (P1, Rehabilitation center)*

> *“Regarding the guidelines of respiratory rehabilitation, there was some difficulty. For example, the use of spirometry was controversial, and most of the respiratory interventions were restricted. With several limitations, there was very little we could actually do, which created problems.” (P11, Government tertiary hospital)*

Physiotherapists stated that fear of infection emerged as a significant concern and impacted in their interaction with the patients which resulted in hesitancy in performing a regular duty shift. They also reported the stress of physical exertion which was associated with working for extended hours.

> *“Everyone working on the frontline during the pandemic including the physiotherapists had fear of infection. As we had to treat our patients, we feared of contracting the infection. Because of that fear, there were a lot of requests like ‘I don’t want to do this shift’, ‘My family is not well’, ‘I think I’m putting my family at risk’, ‘I don’t feel safe enough to come here and work’ and the common feeling among the staff was whether to treat the patients.” (P2, Rehabilitation center)*

### Adaptation, innovation and collaboration should be promoted for strategic intervention and awareness of physiotherapy services during pandemics

#### The change in physiotherapy practice and changing perception

##### Novel approach with Tele-rehabilitation

Physiotherapists stated that they employed a variety of methods to connect with patients, including phone calls, video consultations and tele-therapy sessions. They also reported that videos were frequently used to demonstrate exercises and provide visual guidance, while chats and video calling apps were also frequently used. Some physiotherapists provided hotline numbers to offer immediate support and advice to patients. Patient follow-ups were primarily conducted via phone calls to monitor progress and address patient concerns. One physiotherapist also mentioned when it did not actually work.

> *“…… we contacted the caregiver and showed the video of how to perform the exercises … but he could not deliver that as intended to patient. So, I would say we tried but had to discontinue in several situations as it was difficult.” (P12, Academician)*

##### Change of perception towards physiotherapy

Most of the physiotherapists reported that the pandemic brought about notable shifts in the perception and practice of physiotherapy, with both positive and emerging challenging aspects. There was a marked increase in awareness about the critical role of physiotherapy, particularly in managing respiratory and post-COVID recovery cases. Physiotherapists were increasingly integrated into ICU settings and COVID care teams, where their expertise was highly acknowledged and valued.

> *“………physiotherapists were ignored in the initial days but later, started working in the COVID wards for cardiorespiratory conditions using the WHO protocol. Respiratory interventions were highlighted and also supported by doctors to recover from post-COVID respiratory symptoms …” (P11, Government tertiary hospital)*

#### Strategies for improving and strengthening physiotherapy services during pandemics

##### Strategic mitigation by the physiotherapist or team can resolve the issues during pandemic

The solutions implemented by physiotherapists during the pandemic were aimed at ensuring both the safety of healthcare professionals and the continued provision of essential services to patients. One key solution was the training of staff which helped to better equip them with the necessary skills and knowledge to adapt to the challenges of the pandemic. To manage workloads and maintain patient care, residential duty provisions and modified duty policies were introduced, allowing staff to work in shifts while minimizing exposure. Logistic management was implemented in order to ensure the availability of necessary equipment. Regular communication with administrators provided reassurance and clarity regarding ongoing changes. Protocols for patient care were later developed and regularly discussed to adapt to evolving circumstances. Physiotherapists were encouraged to wear PPE and regularly undergo COVID-19 testing as part of a strategy to protect both staff and patients.

> *“……… due to small number of cases, we started shift wise duties like weekly duty so that many of us do not get exposed at once… We formed two or three groups with one week of duty and two weeks of rest. Along with that, we took precautionary measures too like PPE donning and doffing. At the beginning, we had started the treatment by taking our precautions especially, face protection, eyes protection, which were about wearing masks and gowns.” (P11, Government tertiary hospital)*

##### Patient management strategies are crucial

Physiotherapists reported that they emphasized the importance of proper patient management with treatments being provided only after thorough screening and diagnosis. Patient care was prioritized and treatments were administered based on the urgency and severity of the condition. Measures such as COVID-19 testing, including PCR results and necessary documentation were implemented to ensure safety before treatment. To minimize exposure, modified duty hours and rotational duty hours were introduced. Proper use of protective equipment was encouraged and hand hygiene was properly maintained among physiotherapists and patients as well.

> *“We handled many things related to COVID testing-separating patients, labeling the VTMs (Viral Transport Medium), completing and submitting government forms and then the rehabilitation part. We didn’t collect the actual samples ourselves though.” (P3, Rehabilitation center)*

> *“While entering our department, we didn’t let the patients enter directly. Even for counselling and communication, we maintained proper distance. We didn’t allow any visitors inside without proper sanitization. Washing their hands and wearing masks were compulsorily.” (P5, Government disability sector)*

##### Informing the scope of profession for more recognition

Physiotherapists reported that the pandemic highlighted evolving needs and opportunities in the field. With the growing recognition, it was also mentioned that the priority needs to be that both healthcare professionals and patients should be well-informed about the scope and benefits of rehabilitation services for the better service utilization of physiotherapists.

### Health facility survey

Table 3 outlines the description of health facilities from different districts in province III. The survey included 29 health facilities, 25 in urban and 4 in rural areas and consisted of hospitals (n = 21) and rehabilitation centers (n = 8) which were a mixture of public (37.9%) and non-public (62.1%) settings including private and non-governmental clinics [23].

**Table 3.** Description of the health facilities.

| Districts | Total centers | Tertiary centers | Secondary centers | Medical college centers | Rehabilitation centers |
| --- | --- | --- | --- | --- | --- |
| Kathmandu<sup>a</sup> | 12 | 5 | 3 | 2 | 2 |
| Lalitpur<sup>b</sup> | 5 |  | 1 |  | 4 |
| Bhaktapur<sup>b</sup> | 2 |  | 2 |  |  |
| Kavrepalanchok<sup>b</sup> | 1 |  |  | 1 |  |
| Dolakha<sup>c</sup> | 4 |  | 4 |  |  |
| Makwanpur<sup>b</sup> | 5 |  | 3 |  | 2 |
| Total | 29 | 5 | 13 | 3 | 8 |

<sup>a</sup>Metropolitan<sup>b</sup>Urban, sub-urban<sup>c</sup>Rural district

Table 4 presents the physiotherapy services and role of physiotherapist during the COVID pandemic at various health facilities. More than half of the centers (62.07%, n=18) reported that all physiotherapy services were operational during the pandemic while nearly one third of the centers (31.03%, n=9) reported that the services were halted or not available. PPE was provided in the majority of the centers (89.65%, n=26). Most facilities (86.21%, n=25) reported that physiotherapists worked in a multidisciplinary team. A number of physiotherapists also adopted different roles in fever clinics, COVID receptions, triage and emergency rooms. Others (86.21%, n=25), responded they were involved in physiotherapy services directly handling COVID patients. This involved respiratory interventions, administrative duties and providing support with nursing roles. The majority of the centers (62.07%, n=18) did not implement virtual services at all. It was implemented in some centers (37.93%, n=11). The most common means for virtual care was by phone calls followed by social applications such as Viber, WhatsApp, Zoom, or Skype. Out of the 11 centers that offered virtual care, the service was discontinued in seven. The overall reasons for not implementing the virtual services were related to patients’ accessibility, infrastructure, digital literacy and need for hands on care. Some facilities reported not providing the virtual care, as the physiotherapy services were available at those centers.

**Table 4:** Physiotherapy services and role of physiotherapist at different health facilities (n=29)

| Survey questions | n (%) |
| --- | --- |
| **\*Physiotherapy services during pandemic** |  |
| All services were running | 18 (62.07) |
| OPD patients were treated | 12 (41.38) |
| Involved in direct patient care of those with or at high risk of COVID-19 | 11 (37.93) |
| Acute care services | 10 (34.48) |
| Services were halted | 6 (20.69) |
| Other | 3 (10.34) |
| **Was the PPE easily provided/available to physiotherapist?** |  |
| Yes, easily provided | 23 (79.31) |
| Yes, but there was a shortage. | 3 (10.34) |
| No, lack of PPE | 3 (10.34) |
| **Was the role of Physiotherapist highlighted?** |  |
| Yes-Working in a multidisciplinary team for long term care | 25 (86.21) |
| No | 4 (13.79) |
| **\*Was the role of Physiotherapy different during pandemic?** |  |
| Fever clinic | 6 (20.69) |
| COVID receptions | 4 (13.79) |
| Triage | 4 (13.79) |
| Emergency room posting | 2 (6.90) |
| Others | 25 (86.21) |
| **Did physiotherapist implement Virtual care practice at your center?** |  |
| Not at all | 18 (62.07) |
| Yes, we tried but not continuing now | 7 (24.14) |
| Yes, we are continuing with virtual service | 4 (13.79) |
| **\*Means of Virtual care: n=11** |  |
| Phone call | 10 (90.91) |
| Social applications like Viber, WhatsApp | 2 (18.18) |
| Zoom/Skype | 1 (9.09) |
| **\*Reasons for not providing virtual care: n=18** |  |
| Others | 11 (61.11) |
| Patients unable to receive services | 6 (33.33) |
| Lack of infrastructure | 2 (11.11) |
| Not considered an adequate substitute for hands on care | 2 (11.11) |
| Lack of technical knowledge | 1 (5.56) |

\* Multiple responses

## Discussion

This study provides empirical evidence of how the COVID-19 pandemic significantly impacted physiotherapy services in Nepal, as well as professional role expansion and adaptations. The findings illustrate two realities: substantial interruption of routine physiotherapy services alongside increased visibility of physiotherapy within acute and multidisciplinary care. The discussion here draws on the broader context to understand how these local findings align with global trends and challenges.

Consistent with global reports, our qualitative study revealed variability in physiotherapy services status where a few participants reported uninterrupted services while others shared their experience ranging from compromised service scope and standards to complete shutdown. This is consistent with our quantitative survey results of temporary suspension in nearly one-third of the centers, similar to other studies in India [24], Jordan [25] and Saudi Arabia [26]. WHO revealed that 63% of the member countries reported partial or complete disruption of rehabilitative services [27]. The World Physiotherapy reported that in 2020-2021, 70% of physiotherapy services were disrupted globally, with private practices, public health services and community services being highly affected [2]. Our participants reported a significant decline in patient flow, follow-ups and worsening functional outcomes among patients with disabilities and post-operative needs, echoing global concerns regarding delayed rehabilitation and increased disability burden during the pandemic [28]. The result aligns with a Spanish study where people with chronic health conditions were affected the most due to unavailability of physiotherapy services [29]. The study also revealed that, similar to the findings in Portugal, there was a decline in hands-on treatment during the pandemic [30]. This implied that during a pandemic or similar health crises, physiotherapy services are often undervalued, under-resourced and disrupted, despite a surge in demand for long-term care. The deterioration of patients with chronic conditions, during such events, underscores the longstanding under-integration of physiotherapy within health system planning. The WHO Rehabilitation 2030 initiative emphasizes integrating rehabilitation into all levels of care and strengthening workforce capacity to enhance system resilience [31]. Our study findings also illuminates that there is an urgent need for the proper integration of physiotherapy into emergency preparedness, which is in line with a recent study that showed 83% of countries are not integrating physiotherapy services into national or subnational emergency plans [32,33].

Our study findings on challenges such as lack of transportation or travel restrictions, fear of infection and poor inter-professional collaboration likely contributed to reduced utilization and decreased visits to health facilities, unless absolutely necessary, is consistent with the findings on community members perception and its impact during COVID in Nepal [34]. The findings also revealed that physiotherapists raised concerns about the government initiatives and confusion about the working guidelines. Our study participants (nearly 90%) reported availability of PPE for physiotherapists in contrast to study participants (68%) in Jordan [25], suggesting institutional efforts to sustain essential services and prioritizing safety. However, PPE availability alone did not guarantee continuity, as nearly one-third of facilities suspended services. In addition, our qualitative study revealed that physiotherapists were fearful of getting infected and transmitting infection to their family members and other patients, experienced burn out due to long working hours and found it difficult to work with PPE, consistent with a previous study in Turkey [35].

In order to minimize service interruptions during COVID-19, tele-health quickly spread throughout the world. With 37.93% of centers using virtual care, mostly through phone consultation, tele-rehabilitation emerged as an adaptive response in the Nepali setting, which is line with recent review article on tele-rehabilitation in Nepal [36]. This is also comparable to the patterns observed in studies from India [24,37], Turkey [35] and Kuwait [38], which found that physiotherapists used a variety of techniques, including phone calls and video consultation [39] in addition to applications like WhatsApp and Viber. In contrast, in centers where in-person services persisted, virtual care was almost non-existent, which may suggest that tele-rehabilitation functioned more as a compensatory strategy than a sustained hybrid model. Virtual care was noted as not practical as patients and caregivers had difficulty in implementing the interventions correctly and more hands on services were required. This finding is consistent with the previous studies in Malaysia, Canada, Netherlands, Switzerland and others that stated the barrier to use of tele-rehabilitation was the diminished ability to perform hands-on evaluations [40,41], clinical need for physical contact, technological issues [42,43] and reimbursement frameworks [44].

Importantly, the pandemic created opportunities for recognition and expanded professional roles. The increased recognition of physiotherapy in respiratory and post-COVID recovery care observed in our qualitative study aligns with several studies in Nepal [45,46] and abroad [25,47,48]. Our quantitative results also illuminated that several physiotherapists adapted to different roles and many (86.21%) contributed within multidisciplinary teams for long term care. In order to provide continuous care during the pandemic, physiotherapists in our study emphasized issues such as; their own safety, modifying duty policies, logistic management for necessary equipment and protocols for patient care, including expanding the scope of practice. Our study participants also highlighted that staff training was an important way to deal with the problems during COVID, as also emphasized in other studies for special training [25,48].

These findings reflect broader systemic factors including workforce redeployment, safety and lockdown measures, service prioritization, guidelines, training policies and strengthening digital infrastructure that shaped healthcare delivery during the pandemic. Future preparedness requires policies responding to professionals’ needs to ensure resilience and efficacy in physiotherapy practice amid potential crises [49]. Recognizing the critical role of rehabilitation in pandemic management, the Government of Nepal developed several clinical guidelines to support physiotherapy and rehabilitation services. According to the latest available information, these included: the interim guideline for health-related rehabilitation and physiotherapy of persons with COVID-19 in acute care settings (2020) [50]; the post-COVID-19 rehabilitation management clinical protocol (2022) [51] and protocols related to tele-rehabilitation services. These policy documents indicate national-level recognition and Nepal’s first systematic effort to integrate physiotherapy and rehabilitation into emergency health responses and provide standardized care protocols during a pandemic. However, the extent of awareness, dissemination and implementation of these national guidelines across diverse healthcare facilities remains unclear.

## Limitations of the study

The study was confined to Province III and may not generalize the context of the research findings in different regions of Nepal. The qualitative sample size, although appropriate for thematic analysis, may not capture the full diversity of experiences across different institutional settings.

## Conclusion

The COVID-19 pandemic significantly disrupted physiotherapy services in Nepal while simultaneously highlighting their essential contribution to acute respiratory care, post-COVID rehabilitation and multidisciplinary care. The pandemic exposed systemic issues in healthcare delivery but also demonstrated adaptive capacity within the profession. It emphasized that tele-rehabilitation emerged as a potential alternative for patient care but cannot fully replace in-person services, particularly for manual interventions. It also faced challenges for wider implementation of services due to accessibility issues. Better integration of physiotherapy into national health planning, strengthening rehabilitation infrastructure and institutionalizing hybrid service models are critical to ensuring continuity of patient-centered rehabilitation during future public health emergencies.

## Data Availability

All relevant data are within the manuscript and its Supporting Information files.

## Acknowledgements

We would like to acknowledge the contribution of all study participants, human resources, and logistical support from Norwegian University of Science and Technology, Norway, and Kathmandu University School of Medical Sciences, Dhulikhel Hospital, Nepal. We thank Ms. Ushnis Shakya for assisting in the data collection and Ms. Sulata Karki from Dhulikhel Hospital for the technical support on the data collection toolkit. We would like to acknowledge the contributions of all the stakeholders, directly and indirectly involved, at the federal, provincial, and local levels in public and private settings for facilitating and supporting our study. We are grateful to Ms. Muna Poudel for providing thoughtful comments and suggestions, which helped in improving quality of the manuscript.

## Supporting information

**S1 File. COREQ (Consolidated criteria for reporting qualitative research) checklist**

**S2 File. Interview questionnaire for participants**

**S3 File. Health facility questionnaire for COVID pandemic**

